# Women’s Health And Manifestos (WHAM): UK General Election 2024, a rapid voter information study

**DOI:** 10.1101/2024.06.30.24309732

**Authors:** E Mullins, K Womersley, Fardowsa Abdi, Celestine Donovan-Bradley, Christine Ekechi, Leah Hazard, Jane Hirst, Melanie Nana, Alison Perry, Ana-Catarina Pinho-Gomes, Katherine Ripullone, Stephanie Williams

## Abstract

**Background:** The UK 2024 general election manifestos publicly set out the political parties’ priorities for the eventuality that they are voted into government. We determined to evaluate whether already agreed, evidenced and promoted issues affecting women’s health in the UK had been included in the major parties’ manifestos.

**Methods:** We curated a longlist of priorities and recommendations drawn from major women’s health reports, white papers, national inquiries and health strategies published in the UK over the last 10 years which are publicly available and invited our public contributors to suggest additional topics. We selected the shortlist of women’s health-related priorities – our top 15 ‘asks’ –using a Delphi process. We then devised a scoring system whereby manifestos were marked against the 15 priorities with a maximum of 2 points for each priority. We tested inter-rater reliability on the 2019 Manifesto.

**Results:** Overall, the limited inclusion of prominent issues for women’s health in party manifestos was disappointing across the board. There was little difference between most major parties’ coverage of women’s health in their manifestos. All were limited. Most parties addressed two issues well: childcare and women returning to work after pregnancy; and violence against women and girls and the prosecution of perpetrators.

Several other issues, including assurance that all policy is built in consultation with women; decriminalisation and access to abortion; and women’s health hubs for reproductive, menopause and lifelong health, were considered by none or only one of the major parties.

**Discussion:** Women’s health remains a fringe issue in UK politics, despite the efforts of patients, advocates and healthcare professionals to highlight the suffering that many women live with every day, and at particularly vulnerable and high risk periods of their life such as in pregnancy and the postpartum.

Our analysis highlights the importance of developing previous efforts in women’s health to strengthen existing infrastructure, collaboration and innovation. The next government should build on the work in progress, such as delivering the Women’s Health Strategy (2022) rather than starting afresh.

## Background

The UK 2024 general election manifestos publicly set out the political parties’ priorities for the eventuality that they are voted into government. Manifestos are not binding; they are what the parties promise to do and what the government’s actions will be judged against. By convention, policy commitments included in the manifestos will be supported by all MPs in the ruling party and will not be opposed in the House of Lords.

Amidst the clamour of issues vying for headlines, social media coverage and airtime in personal conversations, we wanted to bring women’s health to the fore of public attention as an issue central to the wealth and wellbeing of the UK. Many organisations and charities publish their own manifestos to put forward their views about what a new government should prioritise. The Women’s Health and Manifestos (WHAM) project group considered that this is to misunderstand the legislative purpose of manifestos and misses an opportunity to scrutinise what is already being pledged.

Instead, we determined to evaluate whether already agreed, evidenced and promoted issues affecting women’s health in the UK had been included in the major parties’ manifestos.

## Methods

The WHAM project brings together a team of people with lived experience in PPIE on women’s health, experts in midwifery, obstetrics and gynaecology, public health, women’s rights, research equity and health inequalities to provide a wide range of perspectives and experience around what matters to women and should be considered urgent to improving their health.

First, the group curated a longlist of priorities and recommendations drawn from major women’s health reports, white papers, national inquiries and health strategies published in the UK over the last 10 years which are publicly available and invited our public contributors to suggest additional topics. These included direct health policies, such as improving access to gynaecology services by reducing the delay for the 591,000 women on gynaecology waiting lists in England alone, as well as more indirect social determinants of health including safe and affordable housing. Our draft shortlist was then reviewed by our public contributors who suggested further priorities with advocacy backing.

We selected the shortlist of women’s health-related priorities – our top 15 ‘asks’ – using a Delphi process. Group members each ranked the longlist to narrow 57 policies and priorities into a shortlist, and then re-ranked iteratively until we agreed on the top 15 priorities. We then devised a scoring system whereby manifestos were marked against the 15 priorities with a maximum of 2 points for each priority. A manifesto was awarded 2 marks for a priority if it was included with emphasis and/or an explicit budget. If a priority was included but in brief or without a funding plan it was awarded 1 point, and if the issue was not mentioned it scored 0. Negative marking was used in response to policies that were deemed by the WHAM group to be regressive for women’s health, with –1 marks given if non-evidence-based or harmful discussion of an issue was included in the manifesto. If raters found positive and negative elements relating to a priority in a manifesto, they allocated an overall score based on an average score. We tested the scoring system’s inter-rater reliability on the 2019 SNP manifesto, and found close agreement in the group’s scoring.

Three group members analysed each of the 2024 manifestos of the major political parties as they were released: Conservative, Labour, Liberal Democrat, Green, Plaid Cymru, SNP, DUP and Reform. An average of the 3 independent scores was taken to produce the overall party manifesto’s score, with the maximum being 30 (2 points for each of the 15 priorities). We then ranked the total scores, with the highest score representing the party who had best recognised the health needs of women in the UK, and the lowest score suggesting that women’s health priorities had not cut through to that party’s manifesto writers or were deemed insufficiently important to include.

Using the scoring data, different group members prepared pitches for outputs in medical journals, newspapers, on social media and other avenues of knowledge sharing. These outputs, along with any impact data available, will be summarised in an update to this paper. The rapid nature of this study, after an unexpected general election announcement and relatively quick campaigning period, had to accommodate the existing commitments of group members. The different release dates of the party manifestos meant that a pragmatic approach was taken at each stage of the project’s development, evaluation of the scoring list and the marking of the manifestos. Deadlines were set for each stage and where these were not feasible for group members to meet, the lead authors, EM and KW, took a joint decision on the validity of each stage and scored the outstanding manifestos.

## Results

Overall, the limited inclusion of prominent issues for women’s health in party manifestos was disappointing across the board (table 1).

**Table 1.** Manifesto scoring for UK 2024 General Election party manifestos.

| Women's Health priority areas | Labour | Liberal Democrat | Conservative | Green | Plaid Cymru | Reform | SNP | DUP | Our rank | Mean manifesto score rank |
| --- | --- | --- | --- | --- | --- | --- | --- | --- | --- | --- |
| Policy and strategy gets built, informed, influenced by women, as a matter of course | — | ✓ | — | ✓ | ✓ | — | — | — | 1 | 6= |
| Ensure healthcare policies address and work towards reducing racial and gender health inequities | ✓ | ✓✓ | — | — | ✓ | — | ✓ | — | 2 | 5 |
| Safe staffing levels in maternity units, address midwifery retention and recruitment crisis | ✓ | ✓ | ✓ | ✓ | ✓ | ✓ | ✓ | — | 3 | 3 |
| Improve access to NHS gynaecology services and address waiting lists | ✓ | ✓ | ✓ | ✓ | — | ✓ | ✓ | ✓ | 4 | 8= |
| Inclusion of women in medical research and clinical trials | — | — | — | — | — | — | — | — | 5 |  |
| Ensuring impact on women of economic policies is considered | ✓ | — | — | ✓ | ✓ | — | — | — | 6 | 6= |
| Decriminalisation of and access to abortion | — | ✓ | — | — | — | — | ✓ | ✗ | 7 | 13 |
| Commit to tackling maternity inequalities, particularly for Black and Asian women | ✓ | — | ✓ | — | — | — | — | — | 8 | 8= |
| Ensure accessibility to safe, secure, liveable and affordable housing for women and their families | — | ✓ | ✓ | ✓ | ✓ | ✓ | ✓ | ✓ | 9 | 4 |
| WH hubs for menstrual, menopause and gynaecological conditions | — | — | ✓ | — | — | — | — | — | 10 | 11= |
| Preconception care, promote optimising health before pregnancy | — | — | — | — | — | — | — | — | 11 |  |
| Equal pay for women, specific measures such as transparent pay reporting, mandatory gender pay gap action plans | ✓ | — | — | ✓ | — | — | ✓ | — | 12 | 11= |
| Improved Postnatal Care and support | — | — | ✓ | — | ✓ | — | — | — | 13 | 10 |
| Wraparound and early years childcare affordable, economic options available to enable women to return to work after pregnancy | ✓ | ✓✓ | ✓✓ | ✓ | ✓✓ | ✗ | ✓✓ | ✓ | 14 | 1= |
| Address unacceptable delays in sexual violence cases in UK and show government priority in addressing VAWG | ✓✓ | ✓✓ | ✓✓ | ✓✓ | ✓✓ | — | — | ✓ | 15 | 1= |
| **Total manifesto score /30** | **10** | **10** | **8.33** | **8.33** | **8.67** | **0.67** | **6.5** | **2.5** |  |  |

***Legend:* ✗***addressed regressively*, **—***not addressed*, **✓***included*, **✓ ✓***included as a policy which was given priority and/or an explicit budget*

A press release was developed for circulation to media outlets which narrates the results section:

***UK political party manifestos ignore key issues in women’s health despite high-profile campaigns and inquiries. Women’s health risks being de-prioritised with a change in government***.

## Main points

- There was little difference between most major parties’ coverage of women’s health in their manifestos. All were limited and scored 7-10 out of a possible 30 points (SNP 7, Labour and Lib Dems 10), while the DUP scored 2.5 and Reform 0.7.
- Most parties addressed two issues well: childcare and women returning to work after pregnancy; and violence against women and girls and the prosecution of perpetrators.
- Two issues – inclusion of women in clinical research and trials, and preconception health – did not feature in any party manifesto which suggests that neither are currently considered matters of urgency in UK politics, in contrast to our group’s opinion.
- Several other issues, including assurance that all policy is built in consultation with women; decriminalisation and access to abortion; and women’s health hubs for reproductive, menopause and lifelong health, were considered by none or only one of the major parties. This highlights a danger that women’s health in the UK could fall off the political agenda and the few gains that have been achieved under the current Conservative Government, most notably the Women’s Health Strategy, risk being undone.
- Although several parties’ general commitments to reducing NHS waiting lists were thought likely to reduce the number of women on gynaecology waiting lists (currently 592,000), our clinical experience is that women with non-cancerous and non-cardiovascular conditions such as endometriosis, prolapse, and heavy menstrual bleeding risk being deprioritised because non-experts may not appreciate the impact that these conditions have on women’s quality of life.
- We recommend that party manifestos and policy platforms would benefit from public and expert input, as is mandated for any proposed UK healthcare or research spending. This would ensure political parties do not ignore or waste the extensive work on identifying unmet needs and recommended actions on women’s health that already exists, including from UK Chief Medical Officers (1), the Women’s Health Strategy in England (2), The Women’s Health Plan in Scotland (3), the Women’s Health Ambassador, the Ockenden(4)/Kirkup(5)/Cumberledge (6) maternity reports/review, and national audits such as MBRRACE (maternal mortality and morbidity).

## Discussion

Overall, the group were concerned and disappointed by the low scores that party manifestos received in our analysis. Women’s health remains a fringe issue in UK politics, despite the efforts of patients, advocates and healthcare professionals to highlight the suffering that many women live with every day, and at particularly vulnerable and high risk periods of their life such as in pregnancy and the postpartum.

The major three parties scored similarly on women’s health, and the scores were not impressive. However, if every priority that were mentioned in the manifestos taken as a group were delivered by the next government, this would significantly improve the lives and livelihoods of women throughout the UK. We also noted a disparity in women’s health priorities across the devolved nations, with SNP and DUP scoring significantly lower than the major three parties as well as Plaid Cymru. Northern Ireland is the only country in the Union to have a pro-life party in mainstream politics and the DUP’s emphatic commitment to restricting access to abortion was negatively marked by our group. The lowest scoring party, Reform, promotes outdated and limiting roles for women, and our analysis highlights how little their ‘contract’ considered women’s health a topic of importance.

Some colleagues who reviewed our analysis questioned why menopause care was not included as a priority. Our group think that this is an area which has benefitted significantly from highly effective campaigning, public support and healthcare response. The conditions in healthcare and in workplaces continue to improve for women experiencing the menopause in response to this advocacy. While complacency would be a mistake, menopause policy progress has been impressive and looks set to continue.

Our analysis also highlights the importance of developing previous efforts in women’s health to strengthen existing infrastructure, collaboration and innovation. The next government should build on the work in progress, such as delivering the Women’s Health Strategy (2022) rather than starting afresh. While it might seem superficially politically expedient to cut ties with this work, the Women’s Health Strategy has brought benefits particularly through its promotion of Women’s Health Hubs to deliver holistic medical care to women, particularly for their cardiac health as well as sexual and reproductive needs.

We also note that our analysis’ 15 priorities have been brought to public attention due to inter-disciplinary approaches within academic settings as well as in collaboration with the public, patients and advocacy groups. All women should have avenues available to influence research agendas. Moreover, researchers should meet their public responsibility by sharing their findings with society to achieve impact and raise awareness.

The WHAM group has found this opportunity to focus on the visibility of healthcare policy rewarding and revealing, and we would commend our approach to other researchers seeking to take the temperature of key issues in UK politics. We intend to reconvene to analyse the priority given to women’s health issues in future election manifestos to ensure women’s health becomes a greater focus for politicians, as it already is for the electorate.

We intend for this analysis to be used as a resource to inspire the next government who may, on reflection, realise they could have done more on women’s health when seeking the electorate’s support. By holding a mirror up to the parties’ underwhelming performance in this election season we hope this might prompt improvement when manifesto writing comes around again, as well as for whichever party starts work in Downing Street next week.

## Data Availability

N/A

